# Community Health Promoter-led responsive feeding for caregivers of children aged 0–59 months in West Pokot, Kenya (CHP-RF-WP Trial): protocol for a pragmatic cluster randomized controlled trial with mixed-methods evaluation

**DOI:** 10.64898/2026.11.22.26368709

**Authors:** Anthony Kipkoech Koross, Everlyne Nyanchera Morema, Silvenus O. Konyole, Morris Senghor Shisanya

## Abstract

**Background:** Responsive feeding — recognising and answering a child’s hunger and satiety cues without coercion — is a World Health Organization-recommended component of complementary feeding, yet it is rarely addressed or measured in Kenya’s community health system, particularly in arid and semi-arid counties.

**Objective:** To determine whether a Community Health Promoter (CHP)-led responsive feeding intervention improves responsive feeding practices among caregivers of children aged 0–59 months in West Pokot County, Kenya, compared with usual community health services, and to evaluate its implementation.

**Methods:** This is a pragmatic, two-arm, parallel-group cluster randomized controlled trial with an embedded RE-AIM mixed-methods process evaluation. Eighteen Community Health Units in two sub-counties will be randomized 1:1, stratified by sub-county, and approximately 340 caregiver–child dyads enrolled. The intervention comprises five-day competency-based CHP training, monthly household counselling and coaching visits over three months, mother-to-mother group sessions, pictorial job aids, SMS reminders and supportive supervision with fidelity assessment. The comparator is usual care. The primary outcome is the Responsive Feeding Practices Index (RFPI, 0–100), combining observed mealtime and caregiver-reported items, at three months after intervention initiation. Secondary outcomes include caregiver knowledge, attitudes and self-efficacy, minimum dietary diversity, meal frequency and age-appropriate feeding. The primary analysis is a baseline-adjusted mixed-effects linear regression with a random intercept for Community Health Unit, by intention to treat. Reach, adoption, fidelity and maintenance, with key informant interviews and focus group discussions, will be integrated by RE-AIM domain.

**Ethics and dissemination:** The trial is approved by the Masinde Muliro University of Science and Technology Institutional Scientific and Ethics Review Committee (MMUST/ISERC/314/2026) and licensed by the National Commission for Science, Technology and Innovation (NACOSTI/P/26/4195285). Findings will be shared through publication, county feedback meetings and a policy brief.

**Trial registration:** Pan African Clinical Trials Registry, PACTR202611787801603; registered 9 November 2026; enrolment planned from 11 January 2027.

## Introduction

Responsive feeding describes the reciprocal exchange in which a caregiver perceives a child’s hunger and satiety signals, responds promptly and contingently, encourages without coercion, and supports developmentally appropriate autonomy at mealtimes. It is one of the behavioural pathways through which the Nurturing Care Framework links caregiving to child growth, self-regulation and cognitive, emotional and social development [1]. The 2023 World Health Organization (WHO) guideline for complementary feeding of infants and young children aged 6–23 months issued a strong recommendation that children in this age band be responsively fed, noting that delivery of the recommendation depends on the capacity of health workers to counsel caregivers and families [2]. Responsive feeding is therefore a guideline-level intervention target with defined delivery requirements.

Systematic review and meta-analysis of parenting interventions delivered in the first three years of life shows improvements in parenting practices, parent–child interaction and child development across low- and middle-income settings [3], and harmonisation work has clarified how responsive feeding recommendations should be integrated into dietary guidance for infants and young children [4]. A mixed-methods study among low-income caregivers of toddlers found responsive feeding practices to be modifiable and associated with better diet quality [5]. Nutrition education and behaviour change communication trials in Ethiopia, including cluster randomized designs delivered through community structures, have improved complementary feeding knowledge and practice [6,7]. However, most trials measure knowledge, dietary diversity or growth, and few prospectively specify a responsive feeding behavioural outcome as the primary endpoint.

In sub-Saharan Africa, where the pooled prevalence of stunting among children aged 6–59 months remains high [8], caregiver feeding style is shaped by workload, household food insecurity and prevailing norms. Observational work in Ethiopia has documented substantial use of pressuring and forced feeding, patterned by caregiver and child characteristics [9]. Comparable findings have been reported in Kenya: a case study in Teso South found responsive feeding difficult to enact under everyday constraints [10], and work in low-income Nairobi settlements documented feeding behaviours that diverge from responsive principles [11].

Kenya delivers community-level promotive and preventive services through Community Health Units (CHUs), each served by a team of Community Health Promoters (CHPs) supervised by Community Health Assistants, under the Primary Health Care Act 2023 and the Kenya Community Health Policy 2020–2030 [12,13]. National survey data continue to show suboptimal infant and young child feeding, with a minority of children aged 6–23 months achieving a minimum acceptable diet [14]. Community platforms have delivered measurable nutrition gains in Kenya: the Baby Friendly Community Initiative improved exclusive breastfeeding [15] and complementary feeding practices [16] in cluster-randomized and quasi-experimental evaluations. Policy review nonetheless indicates that the responsive caregiving domain of the Nurturing Care Framework is the least operationalised in Kenyan community health practice [17]. Responsive feeding counselling is not currently a defined, supervised, competency-assessed CHP task.

West Pokot County is a predominantly arid and semi-arid land (ASAL) county with dispersed pastoral settlement, seasonal food insecurity and variable road access; the county has recorded substantial caseloads of severe and moderate acute malnutrition [18,19]. Local work has identified cultural determinants of malnutrition among children aged 6–59 months in the county [20], and evidence from comparable pastoral ecological zones shows a distinct undernutrition profile relative to agricultural settings [21,22]. These features make West Pokot both a high-need setting and a demanding test of whether a counselling-based intervention can be delivered with adequate reach and fidelity through an existing community platform.

Three gaps remain. First, no randomized evaluation has tested a structured, competency-based, CHP-delivered responsive feeding package integrated into routine Kenyan community health services. Second, responsive feeding is rarely measured as a directly observed behavioural outcome in trials conducted in ASAL settings; measurement typically relies on caregiver report alone. Third, where community nutrition interventions have been evaluated, implementation determinants are seldom captured prospectively alongside effectiveness, so it is not known whether an efficacious package can be delivered at acceptable reach, adoption and fidelity by CHPs carrying existing workloads. Findings from other paediatric populations cannot be extrapolated directly to pastoral ASAL communities, where feeding norms, food availability and household mobility differ [21,22].

This trial therefore adopts an effectiveness–implementation hybrid orientation [23], in which a randomized effectiveness evaluation is the primary aim and a prospectively specified, RE-AIM-guided mixed-methods process evaluation runs concurrently [24,25]. Its objective is to determine whether a CHP-led responsive feeding intervention improves responsive feeding practices, measured by the Responsive Feeding Practices Index (RFPI), among caregivers of children aged 0–59 months in West Pokot County, compared with usual community health services, and to characterise the reach, adoption, implementation fidelity and early maintenance of the package within the county community health system.

## Objectives and hypotheses

### Primary effectiveness objective

To determine the effectiveness of a CHP-led responsive feeding intervention, compared with usual community health services, in improving caregiver responsive feeding practices measured by the RFPI (0–100) at three months after intervention initiation.

### Secondary effectiveness objectives

To determine the effect of the intervention on caregiver responsive feeding knowledge, attitudes and self-efficacy; on minimum dietary diversity among children aged 6–23 months; on age-appropriate meal frequency; on age-appropriate feeding practices among children aged 24–59 months; and on caregiver-reported experience of counselling.

### Descriptive objective

To characterise baseline responsive feeding practices, and their distribution by child age module, caregiver characteristics and household food security, among caregivers of children aged 0–59 months in the participating CHUs.

### Implementation objective

To evaluate implementation of the intervention using the RE-AIM framework, specifically Reach, Adoption, Implementation (fidelity and dose) and early Maintenance.

### Qualitative and contextual objective

To explore barriers to and facilitators of intervention uptake, delivery and sustainability from the perspectives of caregivers, CHPs, supervisors and county health managers.

### Hypotheses

#### Primary hypothesis

Caregiver–child dyads in CHUs allocated to the CHP-led responsive feeding intervention will have higher endline RFPI scores, adjusted for baseline RFPI, than dyads in CHUs allocated to usual care.

#### Secondary hypotheses

Relative to usual care, the intervention will increase caregiver responsive feeding knowledge, attitude and self-efficacy scores; increase the proportion of children aged 6–23 months meeting minimum dietary diversity; and increase the proportion of children receiving age-appropriate meal frequency and age-appropriate feeding practices. Secondary hypotheses are supportive and are not used to establish the primary conclusion of the trial.

## Materials and methods

### Study design and framework

This is a pragmatic, two-arm, parallel-group cluster randomized controlled trial with an embedded mixed-methods effectiveness–implementation (process) evaluation guided by the RE-AIM framework [24,25]. The trial is designed within a superiority framework. The unit of randomization is the CHU; the unit of enrolment, follow-up and analysis is the caregiver–child dyad. The same dyads enrolled at baseline are followed prospectively to the endline assessment. The protocol is reported in accordance with SPIRIT 2025 [26] and the SPIRIT-Children and Adolescents (SPIRIT-C) 2026 extension [27]; the intervention is described following TIDieR principles [28]. Completed SPIRIT 2025 and SPIRIT-C 2026 checklists are provided as S1 File and S2 File. The schedule of enrolment, interventions and assessments is shown in Fig 1, and the trial phases, process activities and division of activities between the arms in Fig 2.

**Fig 1.**
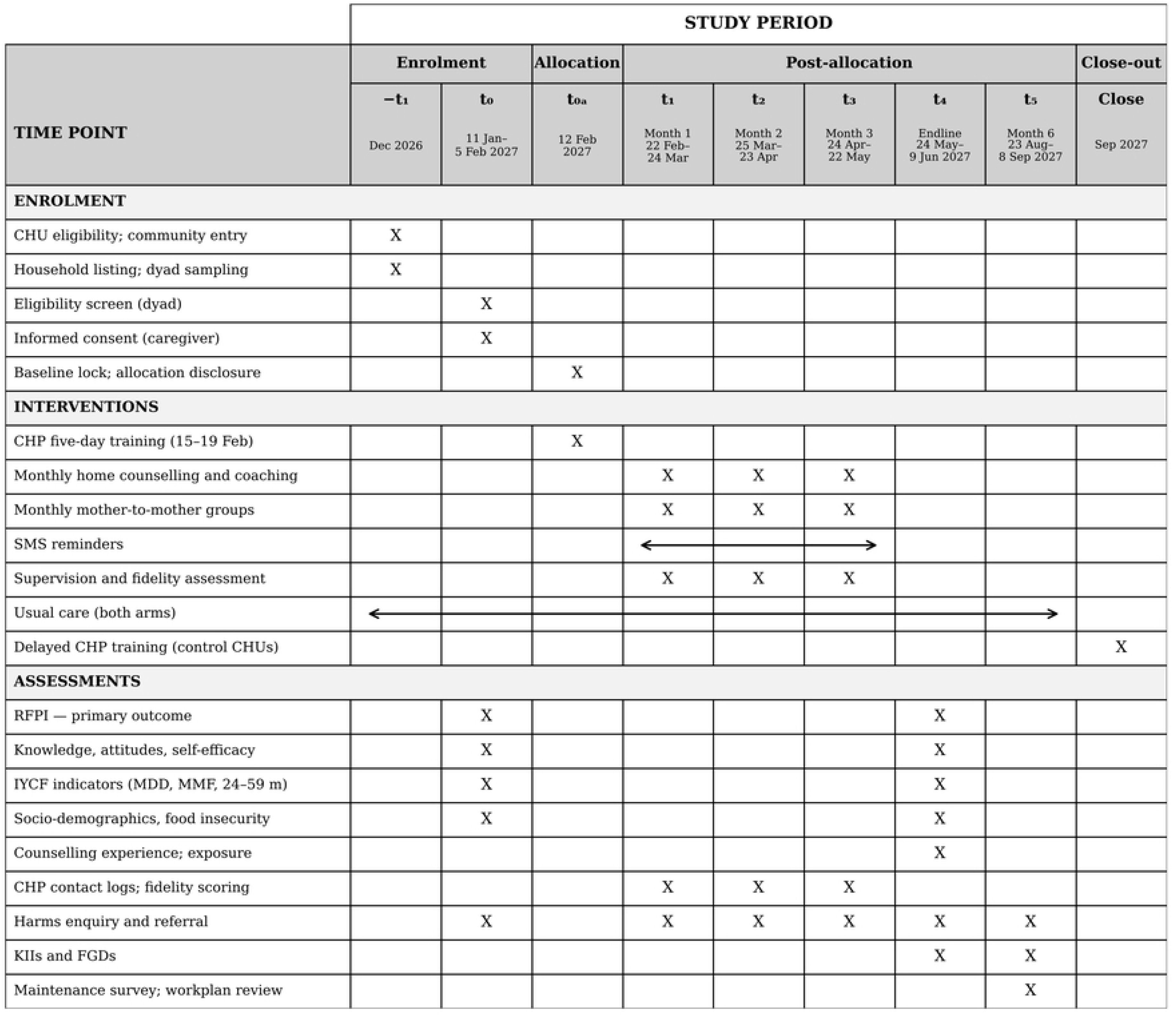
SPIRIT schedule of enrolment, interventions and assessments. X indicates that the activity occurs at that time point; arrows indicate continuous delivery. Allocation is disclosed only after baseline assessment is complete in all 18 CHUs and the baseline dataset is locked; the sequence is generated in advance by an independent statistician and held concealed. The primary effectiveness time point is the month-3 endline; month 6 is an implementation (maintenance) assessment. CHP, Community Health Promoter; CHU, Community Health Unit; IYCF, infant and young child feeding; MDD, minimum dietary diversity; MMF, minimum meal frequency; RFPI, Responsive Feeding Practices Index; SMS, short message service.

**Fig 2.**
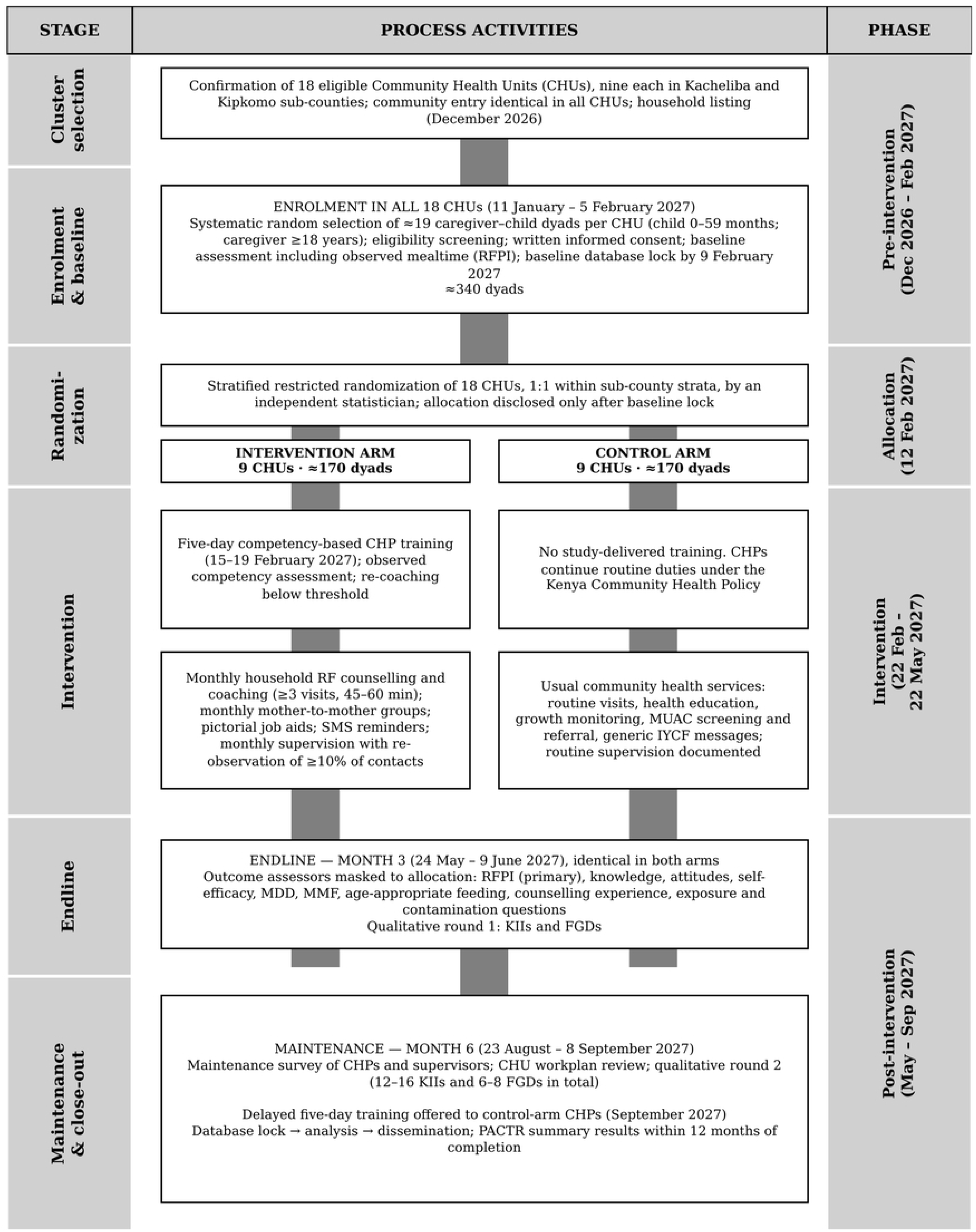
Trial phases, process activities and study arms. Activities shown across both arms are conducted identically in both arms. CHP, Community Health Promoter; CHU, Community Health Unit; FGD, focus group discussion; IYCF, infant and young child feeding; KII, key informant interview; MDD, minimum dietary diversity; MMF, minimum meal frequency; MUAC, mid-upper arm circumference; RF, responsive feeding; RFPI, Responsive Feeding Practices Index; SMS, short message service.

A pragmatic orientation was adopted because the research question concerns whether the package works when delivered by the CHPs who already serve these communities, under existing supervision arrangements and workloads, rather than under idealised conditions. The Nurturing Care Framework provides the content framework, defining responsive caregiving and nutrition as the domains targeted [1]. Orem’s Self-Care Deficit Nursing Theory and Pender’s Health Promotion Model, which underpin the parent doctoral work, articulate the presumed mechanism of change: caregivers experience knowledge, skill and confidence deficits in responsive feeding that CHPs address through counselling and coaching, and adoption of the resulting behaviours is conditioned by perceived benefits, perceived barriers and self-efficacy. RE-AIM is used only as the implementation evaluation framework.

The conceptual framework is presented in Fig 3. The independent variable is the CHP-led responsive feeding package, randomized at CHU level. The intervening variables are caregiver responsive feeding knowledge, attitudes and self-efficacy, which are measured as secondary outcomes but are not treated as established mediators, since the trial is not powered for formal mediation analysis. The dependent variables are the RFPI as the primary outcome and the child feeding indicators as secondary outcomes. Confounding is controlled by cluster randomization, stratification and prespecified covariate adjustment. The RE-AIM layer evaluates whether the intervention was delivered and sustained; it is not part of the causal pathway.

**Fig 3.**
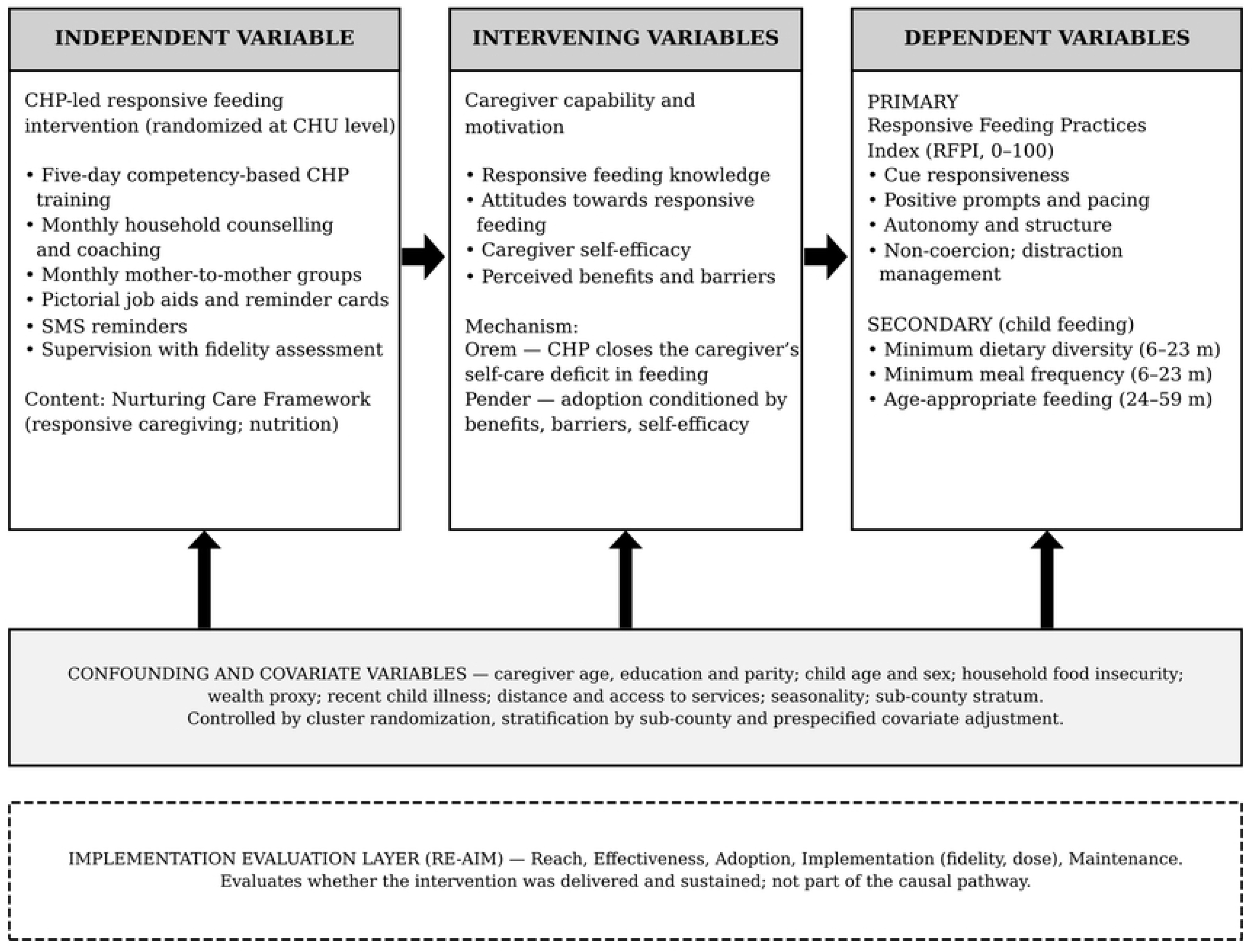
Conceptual framework for the trial. Arrows show the hypothesised pathway from the intervention, through caregiver capability and motivation, to the outcomes. Covariates are controlled by randomization, stratification and prespecified adjustment. The RE-AIM layer evaluates delivery and is not part of the causal pathway. CHP, Community Health Promoter; CHU, Community Health Unit; RE-AIM, Reach, Effectiveness, Adoption, Implementation, Maintenance; RFPI, Responsive Feeding Practices Index; SMS, short message service.

### Trial setting

The trial will be conducted in West Pokot County, north-western Kenya. The county covers approximately 11,075 km² and is administratively divided into six sub-counties (West Pokot, North Pokot, Kacheliba, Pokot Central, Pokot South and Kipkomo), 30 wards and 184 locations, served by 252 CHUs. All sub-counties are classified as arid or semi-arid except Pokot South and part of West Pokot sub-county, which are largely highland. Approximately 80% of the county population resides in ASAL sub-counties, and children under 14 years constitute about half the population [19]. Integrated Food Security Phase Classification analyses have documented substantial burdens of severe and moderate acute malnutrition in the county [18].

The features of this setting that bear directly on the trial are dispersed pastoral settlement, long and seasonally variable travel distances to health facilities, seasonal fluctuation in food availability and household composition, and reliance on the CHP cadre for most household-level promotive contact. These conditions constrain contact frequency, make household visiting the only feasible delivery modality for many families, and create a risk of differential loss to follow-up related to mobility. They also make the findings relevant to other Kenyan and East African ASAL settings.

Two geographically non-adjacent sub-counties, Kacheliba and Kipkomo, were purposively selected as the trial area on the basis of CHU functionality, accessibility for supervision and monitoring throughout the intervention period, and physical separation sufficient to limit cross-arm contact. Sub-county is a stratification and contextual variable only; sub-counties are not randomized and do not define study arms.

### Cluster eligibility

CHUs are eligible for randomization if they are officially gazetted and functional within the county community health information system; have an established CHP team with an assigned Community Health Assistant or equivalent supervisor; have not received a structured responsive feeding or infant and young child feeding counselling programme from any implementing partner in the preceding 12 months; are accessible for monthly supervisory visits throughout the intervention period; and are not immediately geographically adjacent to another CHU already selected for the trial. CHUs are excluded if a partner-implemented nutrition behaviour change programme is scheduled to start in the CHU during the trial period, or if the CHU is subject to insecurity or population displacement that would prevent scheduled household contact. Eighteen eligible CHUs, nine per sub-county, will form the trial sampling frame; where more than nine CHUs are eligible in a sub-county, the nine will be selected to maximise geographical separation, and the selection will be documented before randomization.

### Participant eligibility

#### Inclusion criteria (caregiver–child dyad)

i. Mother or primary caregiver aged 18 years or older who is principally responsible for the index child’s daily feeding; (ii) index child aged 0– 59 months; (iii) resident within the selected CHU catchment for at least three months preceding enrolment; (iv) intention to remain resident within the CHU catchment for the duration of follow-up; (v) written informed consent, including consent for observation of a mealtime.

#### Exclusion criteria

(i) Index child with a condition that fundamentally alters the feeding interaction such that observation of ordinary responsive feeding is not meaningful or not appropriate, including orofacial clefts, severe neurodevelopmental impairment affecting oral intake, and current enteral or tube feeding; (ii) index child currently in inpatient therapeutic care for complicated severe acute malnutrition at screening, since feeding is then clinically directed (such children are referred and may be screened after discharge if follow-up remains feasible); (iii) caregiver unable to provide informed consent. Where more than one eligible child is present in a household, one index child will be selected at random by the electronic data capture form, and only that dyad will be enrolled. No exclusion is applied on the basis of child nutritional status, parity, marital status, socioeconomic position or household composition.

#### Adolescent caregivers

Caregivers younger than 18 years will not be enrolled, because their inclusion would require a separate parental permission and assent framework and additional safeguarding provisions. Adolescent caregivers identified during household listing continue to receive routine community health services, including routine CHP counselling in intervention CHUs, but do not contribute trial data; this limits generalisability to adolescent mothers.

#### Child assent

All index children are aged 0–59 months, so assent is not developmentally applicable and will not be sought. The consenting caregiver gives permission for the child’s participation, including mealtime observation. Child-level safeguards during observation are specified under Harms and safeguarding.

### Community Health Promoter eligibility

CHPs are eligible to deliver the intervention if they are formally registered as CHPs serving a CHU allocated to the intervention arm, are active in the county community health information system, and consent to training, supervision and fidelity assessment. All CHPs serving the nine intervention CHUs will be invited; the CHU establishment in this county is approximately eight to ten CHPs per unit. CHPs in control CHUs continue their routine duties and will be offered the training package after completion of the final trial assessment.

### Recruitment and timing relative to randomization

Recruitment is sequenced so that neither cluster selection nor individual enrolment can be influenced by knowledge of allocation. The following order will be followed and documented:

1. Identification and confirmation of the 18 eligible CHUs against the cluster eligibility criteria, and documentation of the final cluster list.
2. Community entry and sensitisation meetings with sub-county health management teams, CHU committees and community leaders, conducted identically in all 18 CHUs and without reference to arm.
3. Household listing within each CHU to enumerate households containing a resident child aged 0–59 months.
4. Systematic random selection of eligible dyads from the CHU listing, using a random start and a sampling interval derived from the listing size, until the target of approximately 19 dyads per CHU is reached.
5. Individual informed consent.
6. Baseline assessment, including the observed mealtime module where feasible.
7. Disclosure of cluster allocation to the county team, CHPs and field teams, only after baseline data collection has been completed and the baseline dataset for all 18 CHUs has been locked.
8. CHP training in intervention CHUs, followed by initiation of the intervention.

The allocation sequence is generated in advance but concealed until baseline lock (step 7). Enrolment will be tracked against target by CHU, and retention will be supported through collection of two contact telephone numbers, recording of household landmarks, CHP-assisted tracing, appointment reminders through the CHP, and flexible scheduling of endline visits around seasonal movement.

### Randomization and sequence generation

The 18 eligible CHUs will be arranged into two strata corresponding to the two sub-counties, with nine CHUs per stratum. Stratified restricted randomization will be used to achieve nine intervention and nine usual-care CHUs overall. Because each stratum contains an odd number of clusters, the independent statistician will first select at random (probability 0.5) the stratum that receives five intervention CHUs; the other stratum receives four. Within each stratum, the CHU codes will then be placed in a computer-generated random order and the fixed number of intervention CHUs assigned, so that every combination of intervention CHUs within a stratum is equally likely; the remaining CHUs form the usual-care arm. Stratification by sub-county balances the arms on contextual differences in market access, agro-ecology and health service configuration.

The sequence will be generated by an independent statistician who has no role in cluster selection, participant recruitment, intervention delivery, outcome assessment or analysis, using a reproducible pseudo-random number generator in R with a seed recorded before the run. The CHUs in each stratum will be listed in an order fixed before sequence generation and identified to the statistician by anonymised cluster codes only. The sequence, seed and code will be retained by the statistician in a password-protected file and released to the trial team only at the allocation disclosure step.

### Allocation concealment and implementation

The allocation sequence will remain inaccessible to the principal investigator, recruiters, enumerators and CHPs until all baseline procedures across all 18 CHUs are complete and the baseline dataset is locked. Cluster allocation will then be implemented by the independent statistician, who will transmit the arm assignment for each cluster code to the trial coordinator; the coordinator will communicate assignments to the county health management team and to CHU supervisors. No member of the field team will hold the sequence at any time. The date and time of allocation disclosure will be recorded. Because whole clusters are allocated, there is no participant-level allocation step.

### Masking

Masking of CHPs and of caregivers in intervention CHUs is not possible: the intervention consists of counselling and coaching delivered face to face by an identifiable provider. The following masking will be implemented. Endline outcome assessors will be enumerators who were not involved in intervention delivery or supervision, will be assigned to CHUs on a rotating basis where logistics permit, and will not be informed of cluster allocation; caregivers will be asked at the start of each endline visit not to discuss any counselling they may have received until the assessment is complete. The primary analysis will be conducted by the trial statistician, who is separate from the independent statistician who generates and releases the sequence, with arms coded as A and B; the primary analysis will be finalised before arm labels are revealed. The data manager responsible for database lock will be masked to arm labels. Qualitative analysts cannot be masked, because interview content reveals exposure.

Residual risks of observer and ascertainment bias are addressed by using a structured, behaviourally anchored observation instrument with standardised scoring rules, by conducting observer standardisation and inter-rater reliability exercises before and during data collection, by supervisor back-checks on at least 10% of households, and by prespecifying the analysis model. Success of assessor masking will be recorded at the end of each endline visit by asking the assessor to state which arm they believe the cluster was in, and this will be reported.

### Intervention

#### Name and rationale

The intervention is a CHP-led responsive feeding counselling and coaching package grounded in the responsive caregiving and nutrition domains of the Nurturing Care Framework [1] and in the WHO 2023 complementary feeding recommendations on responsive feeding [2]. Its rationale is that caregivers in this setting possess the intent but not consistently the knowledge, skills or confidence to read and answer feeding cues without coercion, and that CHPs are already present in the household and are therefore the appropriate agents to close that gap. Intervention and comparator components are summarised in Table 1.

**Table 1.** Intervention and comparator components.

| Component | Intervention arm (9 CHUs) | Comparator: usual care (9 CHUs) |
|---|---|---|
| Provider | CHPs serving the CHU (approximately 8–10 per CHU), supervised by Community Health Assistants | The same CHP cadre, working to routine job description |
| Provider training | Five-day competency-based responsive feeding training adapted from the UNICEF Community IYCF Counselling Package; end-of-training observed counselling competency assessment with re-coaching for those below threshold | No study-delivered training during the trial; the same five-day training offered after the final trial assessment |
| Theoretical and content basis | Nurturing Care Framework (responsive caregiving and nutrition domains); WHO 2023 complementary feeding recommendation on responsive feeding | Existing national IYCF and community health messaging |
| Core content | Hunger and satiety cue recognition; non-coercive feeding and managing refusal; positive prompting, pacing and positioning; age-appropriate autonomy and self-feeding; dietary diversity, meal frequency and consistency by age; feeding during and after illness; managing feeding difficulties; engaging male partners and household members | Generic health education, growth monitoring and promotion, MUAC screening and referral, immunisation defaulter tracing, general MNCH messaging |
| Individual delivery | Monthly household counselling and coaching visits (minimum three visits over three months, approximately 45–60 minutes each) using an assess–analyse–act sequence with negotiated actions | Routine household visits at the frequency ordinarily practised in the CHU |
| Group delivery | Monthly mother-to-mother support and demonstration sessions at CHU level | None study-delivered |
| Materials | Pictorial counselling cards and flipbook; caregiver take-home reminder card; counselling register; fidelity and supervisory checklists; referral directory | Existing county materials only |
| Digital component | SMS reminders on key practices and upcoming visits, for caregivers who provide a number and consent | None |
| Supervision and fidelity | Monthly supportive supervision with direct re-observation of at least 10% of contacts, fidelity checklist scoring, case review and corrective coaching; benchmark ≥80% mean contact-level fidelity | Routine county supervision only; documented for descriptive comparison |
| Tailoring | By child age module (0–5 versus 6–59 months), by the specific practices needing change in that household, by household food availability, and by preferred language | Not applicable |
| Active duration | Three months of active delivery (22 February – 22 May 2027), followed by maintenance observation to month 6 | Not applicable |
| Concomitant care | All routine county and facility services continue unchanged | All routine county and facility services continue unchanged |
| Discontinuation criteria | Withdrawal of consent; inpatient admission (resumed on discharge); caregiver request to pause; safeguarding or safety concern. Discontinuation does not remove the dyad from follow-up or from the ITT analysis | Not applicable |

CHP, Community Health Promoter; CHU, Community Health Unit; ITT, intention to treat; IYCF, infant and young child feeding; MNCH, maternal, newborn and child health; MUAC, mid-upper arm circumference; SMS, short message service; WHO, World Health Organization.

#### Provider and provider training

CHPs serving the nine intervention CHUs will receive a five-day competency-based training, adapted from the UNICEF Community Infant and Young Child Feeding Counselling Package facilitator materials [29], delivered by the research team with county nutrition and community health staff. The training covers responsive feeding concepts and the Nurturing Care Framework; recognition of hunger and satiety cues in breastfed infants and in older children; non-coercive feeding and management of refusal; positive prompting, pacing and positioning; age-appropriate autonomy and self-feeding; dietary diversity, meal frequency and food consistency by age; feeding during and after illness; management of common feeding difficulties; engaging male partners and other household members; use of pictorial job aids; counselling and negotiation skills; recognition of danger signs and referral; and documentation. Training methods are demonstration, structured role play, supervised practice and case discussion. Competency will be assessed at the end of training using a structured observed counselling checklist with a prespecified pass threshold; CHPs not reaching the threshold will receive targeted re-coaching and repeat assessment before delivering the intervention. This training is distinct from the three-day enumerator training described under Data collection.

#### Materials

Pictorial counselling cards and a flipbook covering cue recognition, non-coercive feeding, responsive positioning and pacing, age-appropriate texture and diversity, and feeding during illness; a household counselling register; a fidelity checklist; a supervisory checklist; and a referral directory. Caregivers receive a take-home pictorial reminder card. SMS reminders reinforcing key practices and signalling upcoming visits are sent to caregivers who provide a functioning telephone number and consent to receive them.

#### Procedures, mode and location of delivery

Delivery is face to face, in the caregiver’s home for individual counselling, and at an agreed community venue for group sessions. Each household visit follows the assess–analyse–act counselling sequence: the CHP observes or discusses a recent feeding episode, identifies one or two specific practices to change, demonstrates or role-models the practice with the caregiver, negotiates a feasible action, and reviews progress at the next visit. Group sessions use practical demonstration of meal preparation and responsive mealtime routines with peer problem-solving.

#### Schedule, intensity and duration

The active intervention period is three months. Each enrolled dyad receives monthly household counselling and coaching visits during this period (a minimum of three visits, each lasting approximately 45–60 minutes), together with monthly mother-to-mother group sessions at CHU level and SMS reminders. Supportive supervision visits with fidelity assessment occur monthly. The active period is followed by a maintenance observation period to six months after initiation, during which no study-delivered intervention is provided and continuation of responsive feeding counselling as routine CHU practice is observed.

#### Tailoring and adaptation

Counselling content is tailored to the index child’s age module (breastfeeding responsiveness for children aged 0–5 months; complementary feeding responsiveness for children aged 6–59 months), to the specific practices identified as needing change in that household, and to household food availability. Counselling is delivered in the language preferred by the caregiver. Adaptations made during delivery will be documented prospectively on the fidelity checklist and reported.

#### Adherence, fidelity and modification

Delivery adherence is monitored through CHP contact registers, monthly supervisory re-observation of at least 10% of counselling contacts, and fidelity checklist scoring (see Intervention fidelity). Where a CHP falls below the fidelity threshold, corrective coaching is delivered and the CHP is re-assessed at the next supervisory visit. There are no participant-level dose-escalation rules.

#### Criteria for discontinuing or modifying the intervention for an individual participant

Counselling will be paused or discontinued for a dyad if the caregiver withdraws consent; if the index child is admitted for inpatient care, in which case counselling resumes after discharge if the family remains resident; if the caregiver or household requests a pause; or if a safeguarding or safety concern arises. Discontinuation of counselling does not remove the dyad from follow-up or from the intention-to-treat analysis, and outcome data will continue to be sought unless the caregiver withdraws consent for further data collection.

#### Concomitant care

All routine county health services, including facility-based nutrition services, integrated management of acute malnutrition, immunisation, growth monitoring and existing health education, continue unchanged and are permitted in both arms. No care is withheld. Prohibited in control CHUs during the trial is the study-delivered responsive feeding package itself, including the training, job aids and SMS component.

### Comparator

The comparator is usual community health service delivery. In control CHUs, CHPs continue their existing scope of work as defined under the Kenya Community Health Policy [13]: routine household visits, general health education, growth monitoring and promotion where scheduled, screening and referral for acute malnutrition using mid-upper arm circumference, immunisation defaulter tracing, and general maternal, newborn and child health messaging including generic infant and young child feeding advice. Control CHUs also retain access to all facility-based services and to any county or partner services available county-wide. What control CHUs do not receive during the trial is the structured five-day responsive feeding competency training, the responsive feeding counselling protocol and job aids, the scheduled monthly responsive feeding coaching visits, the group sessions, the SMS component and the responsive feeding fidelity supervision. Comparator activity is not standardised or intensified for trial purposes, consistent with the pragmatic design, and the actual content and frequency of routine contacts in control CHUs will be documented so that the comparator can be described empirically. On completion of the final trial assessment, CHPs in control CHUs will be offered the same five-day training and job aids, as agreed with the county.

### Contamination prevention and monitoring

Contamination is addressed prospectively through design and monitoring. The two participating sub-counties are geographically separated; immediately adjacent CHUs are not both enrolled; CHPs are instructed in writing not to deliver responsive feeding counselling, job aids or demonstrations outside their assigned CHU, and this instruction is repeated at each supervisory visit; job aids and flipbooks are numbered, issued to named CHPs and accounted for at each supervisory visit; and group sessions are held within CHU boundaries with attendance registers. Movement of CHPs between CHUs, whether through transfer, reassignment or informal assistance, will be recorded. At endline, all caregivers in both arms will be asked the same set of exposure questions covering whether they received responsive feeding counselling, from whom, how often, and whether they received job aids or SMS messages. Cross-cluster contact will also be explored in the qualitative strand. Contamination is reported descriptively by arm and examined in a prespecified sensitivity analysis; the primary analysis remains intention-to-treat by allocated cluster.

### Intervention fidelity

Fidelity is defined prospectively as the percentage of required counselling content correctly delivered during an observed contact, scored against a checklist covering the required modules for the child’s age band: cue recognition, non-coercive feeding, positive prompting and pacing, autonomy and positioning, age-appropriate diversity and frequency, feeding during illness, use of job aids, negotiation of a specific action, and documentation. Each element is scored as delivered, partially delivered or not delivered, and the contact-level fidelity score is the percentage of the maximum attainable score. A contact-level score of at least 80% is the prespecified programmatic benchmark for high fidelity. Fidelity is assessed by supervisors through direct re-observation of at least 10% of household contacts each month and through review of counselling registers for all contacts. Dose delivered (number and duration of contacts per dyad), content coverage (proportion of required modules delivered across the intervention period), participant receipt (caregiver-reported recall of counselling content at endline) and documented adaptations are recorded separately from the fidelity score. Fidelity data are used descriptively, for corrective coaching, and in prespecified secondary and sensitivity analyses; they are not used to redefine the primary analysis population.

### Outcomes

#### Primary outcome

The primary outcome is the RFPI, a continuous composite score ranging from 0 to 100, assessed at three months after intervention initiation (endline). The measurement variable is the RFPI score; the analysis metric is the endline value adjusted for the baseline value; the method of aggregation is the arm mean; and the time point is the three-month endline assessment. Primary and secondary outcomes are summarised in Table 2.

**Table 2.**
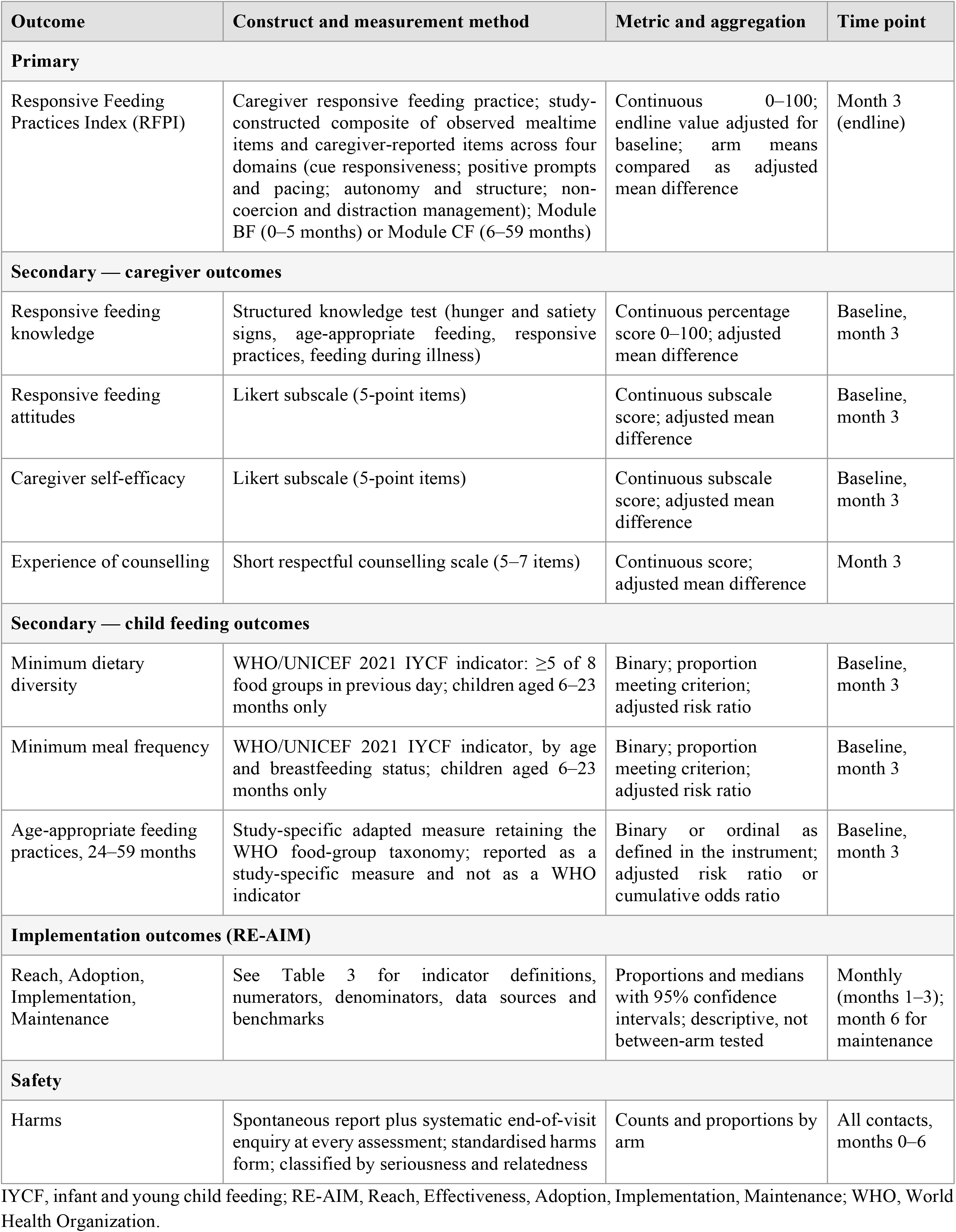
Primary and secondary outcomes, measurement, metric and assessment time points.

| Outcome | Construct and measurement method | Metric and aggregation | Time point |
|---|---|---|---|
| **Primary** |  |  |  |
| Responsive Feeding Practices Index (RFPI) | Caregiver responsive feeding practice; study-constructed composite of observed mealtime items and caregiver-reported items across four domains (cue responsiveness; positive prompts and pacing; autonomy and structure; non-coercion and distraction management); Module BF (0–5 months) or Module CF (6–59 months) | Continuous 0–100; endline value adjusted for baseline; arm means compared as adjusted mean difference | Month 3 (endline) |
| **Secondary — caregiver outcomes** |  |  |  |
| Responsive feeding knowledge | Structured knowledge test (hunger and satiety signs, age-appropriate feeding, responsive practices, feeding during illness) | Continuous percentage score 0–100; adjusted mean difference | Baseline, month 3 |
| Responsive feeding attitudes | Likert subscale (5-point items) | Continuous subscale score; adjusted mean difference | Baseline, month 3 |
| Caregiver self-efficacy | Likert subscale (5-point items) | Continuous subscale score; adjusted mean difference | Baseline, month 3 |
| Experience of counselling | Short respectful counselling scale (5–7 items) | Continuous score; adjusted mean difference | Month 3 |
| **Secondary — child feeding outcomes** |  |  |  |
| Minimum dietary diversity | WHO/UNICEF 2021 IYCF indicator: ≥5 of 8 food groups in previous day; children aged 6–23 months only | Binary; proportion meeting criterion; adjusted risk ratio | Baseline, month 3 |
| Minimum meal frequency | WHO/UNICEF 2021 IYCF indicator, by age and breastfeeding status; children aged 6–23 months only | Binary; proportion meeting criterion; adjusted risk ratio | Baseline, month 3 |
| Age-appropriate feeding practices, 24–59 months | Study-specific adapted measure retaining the WHO food-group taxonomy; reported as a study-specific measure and not as a WHO indicator | Binary or ordinal as defined in the instrument; adjusted risk ratio or cumulative odds ratio | Baseline, month 3 |
| **Implementation outcomes (RE-AIM)** |  |  |  |
| Reach, Adoption, Implementation, Maintenance | See Table 3 for indicator definitions, numerators, denominators, data sources and benchmarks | Proportions and medians with 95% confidence intervals; descriptive, not between-arm tested | Monthly (months 1–3); month 6 for maintenance |
| **Safety** |  |  |  |
| Harms | Spontaneous report plus systematic end-of-visit enquiry at every assessment; standardised harms form; classified by seriousness and relatedness | Counts and proportions by arm | All contacts, months 0–6 |

IYCF, infant and young child feeding; RE-AIM, Reach, Effectiveness, Adoption, Implementation, Maintenance; WHO, World Health Organization.

##### Construction and status of the instrument

The RFPI is a study-constructed composite that has not yet been externally validated. Its observed mealtime module is adapted from published structured observation approaches to caregiver feeding behaviour, and its caregiver-reported module adapts items from existing responsive feeding and feeding practice questionnaires. The composite scoring, item weighting, age modules and missing-data rules are specific to this study and are prespecified here and in the instrument appendix.

##### Domains

Items load onto four conceptual domains: (i) Cue Responsiveness — recognition of and prompt, appropriate response to hunger and satiety cues; (ii) Positive Prompts and Pacing — warm encouragement, modelling and assistance, with pacing matched to the child’s tempo; (iii) Autonomy and Structure — age-appropriate self-feeding opportunity, predictable routine and supportive positioning; (iv) Non-Coercion and Distraction Management — absence of pressure, force, threats or bribes, and management of competing distractions.

##### Age modules

Module BF applies to children aged 0–5 months and assesses breastfeeding responsiveness through five observed items and five caregiver-reported items. Module CF applies to children aged 6–59 months and assesses complementary feeding responsiveness through nine observed items and nine caregiver-reported items, with scoring anchors interpreted against age-appropriate expectations for self-feeding, texture and autonomy. Exactly one module is administered per dyad, determined by the child’s age in completed months at the assessment.

##### Scoring

Observed items are scored 0 (not demonstrated), 1 (inconsistent or partial) or 2 (consistent). Caregiver-reported items use a five-point frequency scale mapped to 0, 0.5, 1.0, 1.5 and 2.0; negatively worded items are reverse-scored before mapping. For Module CF, the observed sum has a maximum of 18 and the reported sum a maximum of 18; RFPI = ((observed sum + reported sum) / 36) × 100. For Module BF, each sum has a maximum of 10; RFPI = ((observed sum + reported sum) / 20) × 100. Domain sub-scores are computed as the percentage of the maximum attainable within the domain. All items carry equal weight. Because both modules are expressed on a common 0–100 metric, scores from the two modules are analysed together, with child age module included as a covariate and as a prespecified exploratory subgroup factor.

##### Missing components

For Module CF, a combined RFPI requires at least seven of nine observed items and at least seven of nine reported items; for Module BF, at least four of five in each set. Where the threshold is met, the sub-score is prorated over the completed items. Where an observed mealtime is not feasible, a reported-only score is computed as (reported sum / maximum reported) × 100, the record is flagged, and the reason is documented. Reported-only records are included in the primary analysis with an indicator variable for observation status, and a prespecified sensitivity analysis is restricted to dyads with a complete observed module.

##### Measurement properties to be assessed

During pretesting (see Pretesting and instrument piloting), internal consistency will be estimated using Cronbach’s alpha and McDonald’s omega for each module and reported as internal consistency estimates, not as evidence of validity. Inter-rater reliability of the observed module will be assessed through paired independent observation of at least 10% of pretest and main-study observations, reported as percentage agreement and as intraclass correlation coefficients or weighted kappa as appropriate. Structural properties will be examined through inter-item and item–total correlations and exploratory factor analysis of baseline data; because the trial is not powered as a psychometric study, these analyses are exploratory. Sensitivity to change will be examined descriptively in the control arm between baseline and endline. Content relevance was reviewed by the supervisory team during instrument development and will be re-examined with county nutrition staff before fieldwork.

#### Observation procedure

Where feasible, one naturally occurring mealtime or snack episode is observed in the home for 10–15 minutes for Module CF, and one breastfeeding episode for Module BF. Caregivers are informed at consent that an ordinary mealtime may be observed and are asked to feed the child as they normally would; no advance coaching or instruction is given, and the observation is scheduled by agreement with the caregiver. The observer remains unobtrusive, does not intervene during scoring except where the child’s safety requires it, records behaviours in real time on the electronic form, and notes start and end times. Observations occur in a location chosen by the caregiver that affords adequate privacy. Enumerators are standardised through video-based scoring drills to a prespecified agreement threshold during training, with repeat drills at mid-point; paired observations for inter-rater reliability are conducted on at least 10% of assessments. If the child becomes distressed, the caregiver wishes to stop, or a safety concern arises, the observation is paused or abandoned and the reason recorded; the dyad then contributes a reported-only score.

#### Secondary outcomes

Caregiver responsive feeding knowledge is measured as a percentage score (0–100) on a structured test covering hunger and satiety signs, age-appropriate feeding, responsive practices and feeding during illness. Caregiver attitudes and self-efficacy are measured using 10–12 five-point Likert items summarised into subscale scores. Minimum dietary diversity is defined per the WHO/UNICEF 2021 indicator guidance as consumption of foods from at least five of eight defined food groups in the previous day, and applies only to children aged 6–23 months [30]. Minimum meal frequency is applied per the same guidance to children aged 6–23 months according to age and breastfeeding status [30]. For children aged 24–59 months, who fall outside the WHO infant and young child feeding indicator age range, a study-specific age-appropriate feeding practice measure retaining the WHO food-group taxonomy will be used and reported separately as a study-specific adapted measure, not as a WHO indicator. Caregiver experience of counselling is measured at endline using a short 5–7 item respectful counselling scale. Household food insecurity is measured using the Household Food Insecurity Access Scale and is treated as a covariate and subgroup factor rather than an outcome [31].

Caregiver knowledge, attitudes and self-efficacy are caregiver behavioural and cognitive outcomes and are reported as such; only the dietary diversity, meal frequency and age-appropriate feeding measures are child feeding outcomes.

#### Implementation outcomes

Implementation outcomes are organised by RE-AIM domain with prespecified numerators, denominators, data sources, measurement frequency and programmatic benchmarks (Table 3). Reach is the proportion of enrolled eligible dyads in intervention CHUs receiving at least one responsive feeding counselling contact, with contact intensity as the mean number of contacts per enrolled dyad. Adoption is the proportion of trained CHPs delivering at least one responsive feeding counselling session, with session volume as the median number of sessions per CHP. Implementation comprises the mean contact-level fidelity score, the proportion of required modules delivered, session completion and documentation completeness. Maintenance is assessed at six months after initiation and comprises the proportion of CHPs and supervisors reporting continued responsive feeding counselling, the proportion of intervention CHUs in which responsive feeding counselling appears in the routine CHU workplan, and reported resource requirements. Effectiveness within RE-AIM is represented by the trial’s primary and secondary effectiveness outcomes.

**Table 3.** RE-AIM implementation indicators, definitions, data sources and prespecified benchmarks.

| Domain | Indicator | Numerator / denominator | Data source | Frequency | Benchmark |
|---|---|---|---|---|---|
| Reach | Coverage of enrolled eligible dyads | N: enrolled dyads in intervention CHUs receiving ≥1 responsive feeding counselling contact / D: all enrolled eligible dyads in intervention CHUs | CHP contact logs; household listing | Monthly; endline | ≥60% of eligible dyads |
| Reach | Contact intensity | N: total counselling contacts delivered / D: enrolled dyads in intervention CHUs | CHP contact logs | Monthly; endline | Mean ≥3 contacts per dyad over the 3-month active period |
| Reach | Representativeness | Comparison of characteristics of dyads reached versus not reached (age, education, food insecurity, distance) | Baseline survey; contact logs | Endline | Descriptive; no threshold |
| Effectiveness | Change in RFPI | Adjusted mean difference in endline RFPI, intervention versus control | Baseline and endline surveys; observation checklist | Baseline; month 3 | ≥6 points (≈0.4 SD) versus control |
| Effectiveness | Minimum dietary diversity (6–23 months) | N: children 6–23 months meeting MDD / D: all children 6–23 months assessed, by arm | 24-hour recall module | Baseline; month 3 | Increase versus control (see Table 2) |
| Adoption | CHP participation | N: trained CHPs delivering ≥1 responsive feeding counselling session / D: CHPs trained | Training register; supervisor reports | Monthly; endline | ≥85% of trained CHPs |
| Adoption | Session volume per CHP | Median number of counselling sessions delivered per CHP over the active period | CHP logs; supervision forms | Monthly; endline | Median ≥6 sessions per CHP over 3 months |
| Adoption | CHU participation | N: intervention CHUs initiating delivery / D: 9 intervention CHUs | Supervision records | Monthly | All 9 CHUs |
| Implementation | Contact-level fidelity | N: fidelity checklist points scored / D: maximum attainable points for the observed contact | Fidelity checklist; supervisory re-observation of ≥10% of contacts | Monthly | ≥80% mean fidelity |
| Implementation | Content coverage | N: required modules delivered across the active period / D: required modules for the child age band | Counselling register | Monthly; endline | ≥80% of required modules |
| Implementation | Documentation completeness | N: contacts with a fully completed register entry / D: all recorded contacts | Counselling register audit | Monthly | ≥90% complete |
| Implementation | Adaptations and deviations | Count and description of documented adaptations; count of protocol deviations by class | Fidelity checklist; deviation log | Monthly | Descriptive; no threshold |
| Implementation | Contamination | N: control-arm caregivers reporting responsive feeding counselling exposure / D: control-arm caregivers assessed at endline | Endline exposure questions (both arms); CHP movement log | Endline | Descriptive; informs sensitivity analysis |
| Maintenance | Continued counselling | N: CHPs and supervisors reporting continued responsive feeding counselling at month 6 / D: CHPs and supervisors surveyed | Maintenance survey | Month 6 | ≥70% |
| Maintenance | Integration in CHU workplan | N: intervention CHUs with responsive feeding counselling in the routine workplan / D: 9 intervention CHUs | CHU workplan review; supervisor interview | Month 6 | Yes in ≥70% of CHUs |
| Maintenance | Resource requirements and local ownership | Reported staff time, materials and supervision needed to continue; qualitative account of ownership | Maintenance survey; KIIs and FGDs | Month 6 | Descriptive; no threshold |

Benchmarks are prespecified programmatic and implementation targets agreed for this trial and its county context; they are not externally validated universal thresholds. CHP, Community Health Promoter; CHU, Community Health Unit; FGD, focus group discussion; KII, key informant interview; MDD, minimum dietary diversity; RFPI, Responsive Feeding Practices Index; SD, standard deviation.

The benchmarks (at least 60% reach, at least 85% CHP adoption, at least 80% mean fidelity and a median of at least six responsive feeding counselling sessions per CHP over the three-month active period) are prespecified programmatic benchmarks agreed for this trial and its county context. They are not externally validated thresholds, and results will be reported against the observed distributions as well as against the benchmarks.

### Process evaluation and qualitative component

#### Purpose and timing

The qualitative strand explains how and why observed implementation and effectiveness patterns arose, and identifies determinants of sustainability. Data collection occurs at two points: at the end of the active intervention period (month 3), and at the maintenance assessment (month 6).

#### Sampling

Purposive sampling with maximum variation will be used. Intervention CHUs will be ranked by a composite implementation performance score derived from reach, adoption and fidelity, and higher- and lower-performing CHUs will both be sampled. Caregivers will be sampled to include those with high and low intervention exposure. CHPs will be sampled to include those with higher and lower fidelity scores. Supervisors and managers will be sampled by role.

#### Participants and volume

Approximately 12–16 key informant interviews will be conducted with county and sub-county health and nutrition managers, CHU in-charges, Community Health Assistants and supervisors, and where relevant partner organisation representatives. Approximately six to eight focus group discussions will be conducted, comprising caregiver groups of 8–10 participants each and CHP groups of 6–10 participants each. These figures are planning estimates; the stopping rule is information power, judged against the specificity of the sample, the strength of dialogue and the analytic aim [32].

#### Procedures

Semi-structured guides organised around Adoption, Implementation and Maintenance, and around barriers and facilitators, will be used. Interviews and discussions will be conducted in the language preferred by participants by trained qualitative researchers who did not deliver the intervention, in a private location, lasting approximately 45–90 minutes. Separate written informed consent, including explicit consent for audio recording, will be obtained. Recordings will be transcribed verbatim, translated into English where required, and back-checked against the recording for a sample of transcripts. Transcripts will be de-identified before analysis, with a separate secure linkage file.

#### Analysis

A hybrid deductive–inductive thematic analysis will follow an established six-phase approach [33]. An initial codebook will be developed from the RE-AIM domains and the interview guides and expanded with inductive codes arising from the data. Two analysts will independently code the first three transcripts to refine the codebook; at least 20% of all transcripts will be double-coded thereafter. Coding discrepancies will be resolved by discussion, with a third team member adjudicating where consensus is not reached. An audit trail of codebook versions, analytic memos and team decisions will be maintained, and the team will record reflexive notes on the influence of professional and institutional positioning on interpretation. Data management and coding will use NVivo or comparable software.

#### Mixed-methods integration

The design is convergent: quantitative and qualitative strands are collected in the same period and integrated at the interpretation and reporting stages, with integration also built in at design level through shared RE-AIM constructs [34]. Integration will be operationalised through joint displays organised by RE-AIM domain, in which quantitative implementation indicators for each CHU are placed alongside corresponding qualitative themes. Convergence, complementarity and discordance will be examined explicitly, and discordant findings will be reported and interpreted. Qualitative findings will also be used to interpret heterogeneity in effectiveness across clusters.

### Participant timeline

The schedule of enrolment, interventions and assessments is presented in Fig 1, and the corresponding phases and process activities in Fig 2. The anticipated calendar is: confirmation of the 18 CHUs, community entry and household listing in December 2026 (month −1); eligibility screening, consent and baseline assessment from 11 January to 5 February 2027 (month 0); baseline database lock by 9 February and allocation disclosure on 12 February 2027; CHP training on 15–19 February 2027; intervention initiation on 22 February 2027, with monthly household counselling contacts, group sessions and supervisory fidelity visits in months 1, 2 and 3 (to 22 May 2027); the month-3 endline effectiveness assessment from 24 May to 9 June 2027; the qualitative strand at months 3 and 6; and the month-6 maintenance assessment from 23 August to 8 September 2027. Close-out, database lock and dissemination follow the maintenance assessment. The maintenance assessment is an implementation assessment and is not a second effectiveness endpoint.

### Sample size

The trial is powered on the primary outcome, the continuous RFPI (0–100). Because no prior RFPI data exist, a baseline mean of 55 and a standard deviation (SD) of 15 are planning assumptions; the pretest will provide a preliminary estimate of variability, but the sample size will not be revised. A between-arm difference of 6 points, equivalent to 0.4 SD, is treated as the minimum programmatically meaningful improvement; it is consistent with the pooled effects of early parenting interventions on parent–child interaction (standardised mean difference 0.39) and parenting practices (0.33) [3].

With a two-sided alpha of 0.05 and 80% power, the individually randomized requirement is n₀ = 2σ²(z₁₋α/₂ + z₁₋β)² / δ² = 2 × 225 × (1.96 + 0.84)² / 36 = 98.1, that is 99 dyads per arm. Approximately 19 dyads will be enrolled per CHU; allowing 10% loss to follow-up leaves about 17 analysable dyads per cluster. With an assumed intra-cluster correlation coefficient (ICC) of 0.03, within the range reported for behavioural and health-related outcomes in community-based studies [35], the design effect is 1 + (17 − 1) × 0.03 = 1.48, giving 99 × 1.48 = 147 analysable dyads per arm. Enrolling approximately 170 dyads per arm (340 in total across 18 CHUs, nine per arm) provides about 153 analysable dyads per arm after 10% attrition.

Because the number of clusters is fixed and small, power was also computed from a non-central t distribution with 2k − 2 = 16 degrees of freedom [36,37]. For an unadjusted comparison of endline means, power is 0.77. The prespecified primary analysis is baseline-adjusted (ANCOVA); assuming a baseline–endline correlation of 0.5, residual variance is reduced by the factor 1 − 0.5² = 0.75 [38], and power is 0.88 (0.86 if loss to follow-up reaches 15%). The cluster-level formula with a t-correction [36] requires eight clusters per arm under ANCOVA (9.5 unadjusted); the planned nine per arm is therefore adequate for the primary baseline-adjusted analysis. A correlation of 0.5 is a conservative assumption for repeated measurement of the same caregiver behaviour three months apart; the observed correlation and ICC will be reported, and no post hoc re-powering will be undertaken.

### Data collection

Data are collected at three levels. Individual (dyad) level: socio-demographic characteristics, caregiver knowledge, attitudes and self-efficacy, RFPI observed and reported modules, infant and young child feeding indicators from a 24-hour recall, child illness in the preceding two weeks, household food insecurity, distance and access, and, at endline, exposure questions and the counselling experience scale. CHP level: training attendance and competency assessment results, contact registers, session records, fidelity checklist scores and supervisory notes. Cluster level: CHU characteristics, CHP establishment and staffing changes, group session registers, supervisory visit records, workplan review at maintenance, and documented contamination events.

Baseline and endline surveys use the same instruments and procedures in both arms. Enumerators and research assistants will receive a three-day training covering research ethics and consent, the study instruments, tablet-based data capture, interviewing technique, observation scoring drills and referral procedures, followed by supervised field practice and an inter-rater reliability exercise.

#### Electronic data capture

Data are captured electronically using KoboToolbox forms on mobile devices, with skip logic, required fields, range and logical consistency checks, constrained response options, automatic timestamping of form start and end, and daily encrypted synchronisation to a secure server. Access is role-based, with distinct permissions for enumerators, supervisors, the data manager and the statistician. Devices are password-protected and forms are not retained locally after successful synchronisation. Global positioning system coordinates will be captured only for household re-location at endline and only where the caregiver consents.

#### Pretesting and instrument piloting

The study instruments, including the RFPI observed and reported modules, will be pretested in Tiaty West Sub-County, Baringo County, in December 2026. This is an instrument pretest and observer standardisation exercise, not a pilot randomized trial, and no randomization, allocation or intervention delivery occurs there. Tiaty West was selected because it is an ASAL sub-county with predominantly pastoral livelihoods and a malnutrition burden comparable to the trial area, and because it lies outside West Pokot County and therefore outside the sampling frame. The pretest will assess comprehension and cultural appropriateness of items, administration time, feasibility of the observed module, internal consistency and inter-rater reliability, and will inform final item wording and enumerator training. Pretest participants are not enrolled in the trial and pretest data will not be included in any trial analysis.

#### Quality assurance

Quality assurance procedures comprise standardised enumerator training with a certification threshold; observer standardisation drills and repeat inter-rater reliability exercises; daily field supervision and end-of-day team debriefs; supervisor back-checks by re-visit or telephone verification in at least 10% of households; automated completeness and consistency reports generated weekly; review of records flagged for extreme values, high rates of not-applicable responses or implausibly short administration times; documented correction procedures with all changes captured in the audit trail; and monthly data quality reports to the trial coordinator.

### Data management

Each cluster receives a unique CHU identifier and each dyad a unique participant identifier constructed from the cluster code and a serial number. Identifying information (names, telephone numbers, household locators) is stored in a separate, access-restricted linkage file and is never held in the analysis dataset. All data are stored encrypted at rest on a secure institutional server with restricted, role-based access and automated daily backup to a separate encrypted location. Datasets are versioned, and a data dictionary and codebook are maintained alongside each version. Analysis datasets are derived from locked source data by documented scripts. The database will be locked after completion of the maintenance assessment and resolution of all data queries; unlocking thereafter requires written authorisation from the principal investigator and is recorded. All trial investigators named on this protocol will have access to the final analysis dataset. Study data will be retained for at least five years after completion in accordance with institutional policy and the Kenya Data Protection Act, 2019.

### Statistical methods

#### Analysis populations

The primary analysis follows the intention-to-treat principle appropriate to a cluster randomized trial: every enrolled dyad is analysed according to the arm to which its CHU was randomized, irrespective of the number of counselling contacts received, CHP fidelity or any cross-arm exposure. Clusters are not excluded after randomization. A per-protocol population, defined as dyads in intervention CHUs receiving at least three counselling contacts at a contact-level fidelity of at least 80%, together with all control dyads reporting no responsive feeding counselling exposure, will be used only for supportive analysis. The analysis plan is summarised in Table 4.

**Table 4.** Statistical analysis plan by objective and outcome.

| Objective | Outcome or variable | Data type | Analysis method | Effect measure reported |
|---|---|---|---|---|
| Descriptive: baseline responsive feeding practices | RFPI total and domain sub-scores at baseline | Continuous | Descriptive statistics by arm, CHU, age module and caregiver characteristics; no hypothesis testing of baseline balance | Means, SD, proportions; ICC estimate |
| Primary effectiveness | RFPI at month 3 | Continuous 0–100 | Mixed-effects linear regression (ANCOVA), random intercept for CHU, fixed effect for arm, adjusted for baseline RFPI, stratum, age module, child age, child sex, caregiver education, household food insecurity; Kenward–Roger degrees of freedom; ITT by allocated cluster | Adjusted mean difference with 95% CI and p-value; standardised effect size; ICC with 95% CI |
| Secondary effectiveness | Knowledge, attitude, self-efficacy, counselling experience scores | Continuous | Mixed-effects linear regression with random intercept for CHU and the same covariate set, adjusted for baseline where measured | Adjusted mean difference with 95% CI |
| Secondary effectiveness | Minimum dietary diversity; minimum meal frequency | Binary | Mixed-effects log-binomial regression; modified Poisson with robust cluster variance if non-convergent | Adjusted risk ratio with 95% CI |
| Secondary effectiveness | Age-appropriate feeding practices (24–59 months); ordinal Likert-derived measures | Binary or ordinal | Mixed-effects log-binomial regression, or cumulative link mixed model with proportional odds assessment | Adjusted risk ratio or cumulative odds ratio with 95% CI |
| Implementation (RE-AIM) | Reach, adoption, fidelity, dose, coverage, maintenance | Proportions, counts, medians | Descriptive summary with prespecified numerators and denominators (Table 3); cluster-level and CHP-level summaries; no between-arm testing | Proportions and medians with 95% CI; comparison against benchmarks |
| Qualitative and contextual | Barriers, facilitators, sustainability determinants | Textual | Hybrid deductive (RE-AIM) and inductive thematic analysis; ≥20% double coding; audit trail; reflexivity notes | Themes with illustrative anonymised data extracts |
| Integration | Convergence between implementation indicators and themes | Mixed | Convergent design; joint displays by RE-AIM domain; examination of convergence, complementarity and discordance | Joint display matrices; narrative integration |
| Supportive | Sensitivity and subgroup analyses | As above | Observed-only versus reported-only RFPI; alternative scoring; contamination-informed; cluster-size weighting and cluster-level analysis; deviation exclusion; per-protocol; missing-data assumptions; treatment-by-subgroup interaction tests | Effect estimates with 95% CI; interaction p-values labelled exploratory |
| Safety | Harms | Counts | Descriptive tabulation by arm and by seriousness and relatedness | Counts and proportions |

ANCOVA, analysis of covariance; CI, confidence interval; CHP, Community Health Promoter; CHU, Community Health Unit; ICC, intra-cluster correlation coefficient; ITT, intention to treat; RFPI, Responsive Feeding Practices Index; SD, standard deviation.

#### Primary analysis

The primary analysis is a mixed-effects linear regression of endline RFPI with a random intercept for CHU, a fixed effect for allocated arm, adjustment for baseline RFPI, and adjustment for the prespecified covariates listed below. This analysis-of-covariance formulation is more efficient than analysis of change scores when baseline and endline are measured on the same dyads and avoids regression-to-the-mean artefacts. The treatment effect is reported as the adjusted mean difference in RFPI with a 95% confidence interval and a two-sided p-value at alpha 0.05, together with a standardised effect size and the estimated ICC with a confidence interval. Denominator degrees of freedom will be based on the number of clusters using the Kenward–Roger approximation. Model assumptions will be examined through inspection of residuals at both levels; if the residual distribution is materially non-normal, a cluster-level bootstrap or a rank-based alternative will be used and reported.

#### Prespecified covariates

The following covariates are prespecified and are included irrespective of their statistical significance: baseline RFPI; randomization stratum (sub-county); child age module (0–5 versus 6–59 months); child age in months; child sex; caregiver education; and household food insecurity. Covariates are not selected using univariable p-value screening. The covariate set is deliberately limited because the trial has 18 clusters; no cluster-level covariate other than the stratification factor is included in the primary model.

#### Secondary outcome analyses

Continuous secondary outcomes (knowledge, attitude, self-efficacy and counselling experience scores) are analysed using mixed-effects linear regression with a random intercept for CHU and the same covariate set, with adjustment for the corresponding baseline value where measured. Binary secondary outcomes (minimum dietary diversity among children aged 6–23 months; minimum meal frequency) are analysed using mixed-effects log-binomial regression to obtain risk ratios; where the model fails to converge, modified Poisson regression with robust cluster-level variance estimation will be used [39]. Risk ratios rather than odds ratios are reported because these outcomes are common. Ordinal outcomes, where they arise from Likert-based items analysed at item level, are analysed using cumulative link mixed models with a random intercept for CHU; the proportional odds assumption will be assessed, and a partial proportional odds or robust alternative used where it is violated. Implementation outcomes are summarised descriptively with cluster-level and CHP-level denominators and 95% confidence intervals; they are not subjected to between-arm hypothesis testing.

#### Multiplicity

One primary outcome and one primary hypothesis are specified, and no adjustment for multiplicity is applied to it. Secondary outcomes are interpreted as a set, primarily through effect sizes and confidence intervals; the number of comparisons will be stated and secondary findings described as supportive. Subgroup analyses are labelled exploratory. No interim efficacy analysis is planned.

#### Missing data

The extent, pattern and stated reasons for missing data will be tabulated by arm, distinguishing item-level missingness, missing observed modules and complete loss to follow-up. Baseline characteristics of dyads retained and lost will be compared descriptively. Dyads lost to follow-up will not be replaced; the cohort is fixed at baseline. The primary model, being likelihood-based, provides valid inference under a missing-at-random assumption conditional on the covariates and the observed baseline value. Where endline outcome data are missing for more than 10% of dyads, multiple imputation by chained equations will be used as a supportive analysis, with the imputation model including arm, baseline RFPI, all analysis covariates and cluster membership, so that it respects the clustered structure and is congenial with the analysis model. Departures from missing-at-random will be examined through a tipping-point analysis applying successively less favourable assumptions to missing intervention-arm outcomes. Missing values of prespecified baseline covariates will be handled by single imputation with a missing-indicator, an approach that is valid for baseline covariates in randomized trials [40], so that every prespecified covariate is retained in the primary model.

#### Sensitivity and subgroup analyses

Prespecified sensitivity analyses are: (i) restriction to dyads with a complete observed RFPI module; (ii) alternative RFPI scoring, using domain-mean scoring in place of item-sum scoring; (iii) exclusion of control dyads reporting substantive responsive feeding counselling exposure; (iv) weighting for cluster-size imbalance, and a cluster-level (two-stage) analysis of cluster mean scores; (v) exclusion of dyads with major protocol deviations; (vi) a per-protocol analysis defined by fidelity and dose as above; and (vii) alternative missing-data assumptions as described. None of these replaces the primary intention-to-treat analysis; all are reported alongside it.

Prespecified exploratory subgroup analyses examine effect modification by child age module (0–5 versus 6–59 months), household food insecurity status, randomization stratum and baseline RFPI tertile, using treatment-by-subgroup interaction tests. Subgroup findings will be interpreted as hypothesis-generating; the trial is not powered for interaction. Analyses will be conducted in R or Stata. The full statistical analysis plan will be finalised and dated before the endline database lock and will be made available in the trial registry record.

### Harms and safeguarding

This is a low-risk behavioural intervention delivered within routine community health services, and no physical or pharmacological risk is anticipated. Foreseeable harms are nonetheless specified. Participant burden arises from survey administration (approximately 45–60 minutes) and mealtime observation; discomfort or self-consciousness may arise during observation; social discomfort may arise if counselling is perceived as criticism of a caregiver’s feeding; confidentiality risks arise from household-level data collection in small communities; and inappropriate advice could be given by a CHP whose competence has drifted, which the competency assessment, fidelity monitoring and corrective coaching are designed to prevent. Counselling could also unintentionally increase caregiver anxiety about a child’s intake; counselling protocols therefore emphasise non-blaming, strengths-based framing, and CHPs are trained not to set intake targets.

Harms will be assessed through spontaneous report to CHPs, supervisors or enumerators, and systematically through a standard end-of-visit enquiry at every household assessment. All reports are recorded on a standardised form, reviewed by the principal investigator within 72 hours, and classified by seriousness and relatedness to trial participation. Serious events, defined as any event resulting in hospitalisation, significant disability or death of a participant or index child, and any safeguarding concern, are reported to the principal investigator within 24 hours and to the Masinde Muliro University of Science and Technology Institutional Scientific and Ethics Review Committee (MMUST-ISERC) within 72 hours of notification, and to the county health authority. A cumulative harms log is maintained and will be reported by arm.

#### Referral

Any index child identified during any study contact with a mid-upper arm circumference or other indicator meeting county thresholds for moderate or severe acute malnutrition, with nutritional oedema, with a persistent feeding difficulty, or with danger signs of serious illness, is referred immediately to the nearest health facility using the county referral protocol, in both arms; referral is never delayed for study procedures. Referrals are documented and followed up by the CHP, and referral completion is recorded.

#### Child safeguarding

SPIRIT-C safeguards are applied [27]. Observation is limited to a single naturally occurring feeding episode of 10–15 minutes. Feeding is not interrupted or manipulated for study purposes. If the child becomes distressed, the observation is paused or ended. The caregiver may stop the observation at any time without giving a reason and without any effect on services received. Observation takes place in a private setting chosen by the caregiver. No photographs or video recordings of children are taken. Children are never left alone with a study team member or CHP. Any suspicion of child harm or neglect is escalated to the principal investigator and referred to the county child protection authority in line with Kenyan law, and this is stated in the consent process. Study staff and CHPs receive safeguarding orientation as part of training, and CHPs are trained not to label ordinary refusal or fussiness as a disorder. Participation does not alter the child’s daily routine: assessments take place at home at times chosen by the caregiver, and the only child-facing procedure is observation of one ordinary feeding episode. Families will be thanked individually at the endline visit and recognised collectively at the community feedback meetings; no payment is made for participation. No assessment of outcomes or harms is planned after the month-6 maintenance assessment; children continue to receive routine county health and nutrition services, including referral, after the trial.

### Monitoring

#### Data monitoring committee

A formal independent data monitoring committee is not constituted. The intervention is a behavioural counselling package delivered within routine services, carries no anticipated serious risk, has a short follow-up period, and includes no interim efficacy analysis and no stopping rules. Safety and conduct oversight is exercised by the trial oversight arrangement described below, and any serious concern is escalated to the ethics committee, which retains the authority to require modification or suspension of the trial.

#### Trial monitoring

Trial conduct is monitored monthly. The trial coordinator reviews recruitment and retention against target by CHU, contact and fidelity records, data completeness and quality reports, protocol deviations and the harms log. Protocol deviations are recorded on a standard form, classified as minor or major, and reviewed by the principal investigator; major deviations are reported to the ethics committee. The doctoral supervisory team, comprising the co-authors of this protocol, reviews progress quarterly and reviews the deviation and harms logs. Monitoring is conducted by the trial team; there is no external monitoring or audit function.

### Patient and public involvement

County and sub-county health managers and CHU committees were consulted during protocol development on feasibility, contact frequency and acceptability of mealtime observation, and their input informed the visit schedule and the decision to schedule observation by agreement with caregivers. During the instrument pretest, caregivers will be asked directly about the comprehensibility, acceptability and intrusiveness of the RFPI items and the observation procedure, and item wording will be revised in response. CHU committees and community leaders will be engaged at community entry in all 18 CHUs, consistent with recommendations for beneficiary and stakeholder participation in community-based nutrition interventions [41]. Caregivers and CHPs contribute to interpretation of findings through the qualitative strand and through community feedback meetings held before final reporting. Participants were not involved in the original formulation of the research question, which arose from the doctoral candidate’s prior work in the county. Plain-language summaries will be provided to participating communities.

### Research ethics approval

Ethical approval has been granted by MMUST-ISERC, which is accredited by the National Commission for Science, Technology and Innovation (NACOSTI) to review research proposals on its behalf (approval MMUST/ISERC/314/2026, 17 August 2026, valid to 17 August 2027). A research licence has been issued by NACOSTI (licence NACOSTI/P/26/4195285, issued 14 September 2026, valid to 14 September 2027, covering Baringo and West Pokot Counties). Because the maintenance assessment runs to September 2027, renewal of the ethics approval and the research licence will be sought before they expire. Written authorisation will be obtained from the West Pokot County Department of Health and its research committee, and from the Kacheliba and Kipkomo sub-county health management teams. The trial will be conducted in accordance with the Declaration of Helsinki [42] and applicable Kenyan law, including data protection requirements. No participant will be enrolled before all approvals are in place.

### Consent

Written informed consent will be obtained from every caregiver before any study procedure, by trained research assistants, in a private setting and in the language the caregiver prefers, using an information sheet and consent form approved by the ethics committee. The consent process covers the purpose of the study, what participation involves at baseline, endline and maintenance, that a mealtime may be observed, the expected time commitment, the absence of individual financial benefit, foreseeable risks and discomforts, the referral pathway, confidentiality and its limits including mandatory safeguarding reporting, the voluntary nature of participation, the right to withdraw at any time without any effect on services received, data storage and retention, and the plan for future data sharing. Adequate time is given for questions. Where a caregiver cannot read, the information sheet is read aloud in the presence of an impartial witness who countersigns, and the caregiver provides a thumbprint. Caregivers provide permission for the index child’s participation. The information sheet and consent form are provided in English, Kiswahili and Pokot; the ethics-approved English version is included in S3 File and the translated versions are available on request.

Separate written informed consent, including specific consent for audio recording, is obtained for key informant interviews and focus group discussions. CHPs provide separate written consent for training participation, fidelity observation and use of their delivery records for research purposes; refusal carries no consequence for their position or duties. Consent for use of de-identified individual participant data in future secondary analyses and for controlled data sharing is sought explicitly within the main consent form, and a caregiver may participate in the trial while declining future data sharing. No biological specimens are collected.

### Confidentiality

Personal information is collected only where required for eligibility, follow-up and referral. Names, telephone numbers and household locators are recorded in a linkage file held separately from the research data, accessible only to the trial coordinator and data manager, and are not exported into analysis datasets. All research records use participant and cluster identifiers only. Paper consent forms are stored in a locked cabinet at the study office, separately from data. Electronic data are encrypted in transit and at rest, and access is role-based. Members of the research team sign confidentiality undertakings. In the qualitative strand, transcripts are de-identified and potentially identifying details are masked or generalised before analysis; because the number of senior county informants is small, quotations will be attributed only by broad role category. Focus group participants are asked to agree at the outset not to repeat what others say, and are informed that confidentiality within a group cannot be guaranteed. Data will be retained for at least five years after completion, after which identifiers will be destroyed.

### Protocol amendments

Substantive amendments to the protocol, defined as changes to eligibility, outcomes, sample size, randomization, the intervention, the analysis plan or the consent process, require the approval of the principal investigator and the supervisory team, and will be submitted for approval to MMUST-ISERC and, where required, to NACOSTI and the county health authority, before implementation. Approved amendments will be reflected in the PACTR record, communicated in writing to all participating CHUs, supervisors, CHPs and research team members, and incorporated into a revised protocol version with an incremented version number and date. Where an amendment affects information material to a participant’s decision to continue, participants will be re-consented. Amendments will be reported in the trial publications.

### Dissemination

Findings will be disseminated regardless of the direction of the results. Planned outputs are peer-reviewed publication of the main effectiveness results, reported in accordance with CONSORT 2025 and its extensions for cluster randomized trials and for outcomes [43–45], of the process evaluation and of the RFPI measurement work; posting of summary results in the PACTR record within 12 months of trial completion; a policy brief for the West Pokot County Department of Health and the national Division of Community Health Services; feedback meetings in participating CHUs at which plain-language summaries are presented to caregivers, CHPs and community leaders; and presentation at national and regional scientific meetings. Plain-language summaries will be provided in the languages used in the study communities. Authorship will follow the International Committee of Medical Journal Editors criteria; no professional medical writers will be used, and no funder or sponsor imposes publication restrictions.

### Data availability and data sharing

No datasets were generated or analysed for this protocol. On completion of the trial, the de-identified individual participant dataset underlying the published results, together with the data dictionary and codebook, will be deposited in a public repository (Zenodo) under controlled access, with requests handled by a non-author data access contact at MMUST-ISERC, subject to approval of a written analysis proposal and a data use agreement. The statistical analysis code and the intervention materials, including the training curriculum, counselling job aids, fidelity checklist and the RFPI instrument with its scoring key, will be made openly available without restriction. Qualitative transcripts will not be shared openly, because the informant pool includes a small number of identifiable county and sub-county roles; requests for qualitative data will be considered individually under controlled access, with ethics committee approval. This plan is conditional on the consent obtained and on ethics committee approval, and is identical to the individual participant data sharing statement in the trial registry record.

### Study status

Protocol version 1.1, dated 15 November 2026 (version 1.1 revises the anticipated dates of version 1.0, 30 September 2026). The trial was registered with the Pan African Clinical Trials Registry on 9 November 2026 (PACTR202611787801603), before enrolment of any participant. Recruitment status at the version date: not yet recruiting; no participant has been enrolled and no trial data have been collected. Anticipated first enrolment: 11 January 2027 (enrolment and baseline 11 January – 5 February 2027; allocation disclosure 12 February 2027; intervention initiation 22 February 2027). Anticipated primary completion (month-3 endline): 9 June 2027. Anticipated study completion (month-6 maintenance assessment): 8 September 2027. The PACTR record was updated on 15 November 2026 to match these dates and the design details of this protocol; the update is under registry review.

## Discussion

This trial addresses a specific gap: responsive feeding has become a guideline-level recommendation [2] without a corresponding evidence base on how it can be delivered through the frontline cadres that reach households in low-resource, dispersed settings. By randomizing CHUs rather than individuals or sub-counties, the design tests the package as it would be deployed — through an intact community health team, under existing supervision, within an existing workplan — while preserving the internal validity of a randomized comparison.

The trial prospectively specifies a behavioural responsive feeding outcome as its primary endpoint, rather than treating responsive feeding as a mediator of dietary or growth outcomes. Measurement combines direct observation of feeding interaction with caregiver report, which addresses the susceptibility of self-reported feeding style to social desirability. Because reach, adoption, fidelity and maintenance are prespecified with defined numerators and denominators, the trial will also produce the information a county health management team would need to decide whether to adopt the package [23–25].

Delivering counselling through CHPs with existing caseloads, without supplementing staffing, means the estimated effect is an effect under realistic conditions and is likely to be smaller than an efficacy estimate obtained under enhanced supervision. The hybrid orientation means that a modest or null effect accompanied by low fidelity would remain interpretable, because the process data will distinguish failure of the intervention concept from failure of its delivery.

For ASAL settings specifically, the design tests whether a home-visiting behavioural intervention is feasible where households are dispersed and seasonally mobile. Retention, contact frequency and observation feasibility in this context are themselves findings of practical value. Because the intervention uses existing cadres, existing supervision structures and low-cost materials, a positive result could inform scale-up through the national community health platform [13], and the maintenance assessment provides an early indication of whether integration into CHU workplans occurs without external support.

Several limitations are anticipated. Blinding of CHPs and of caregivers receiving counselling is impossible, and although endline assessors and the analyst are masked, reporting bias in caregiver-reported items remains possible; the observed module and the prespecified sensitivity analysis comparing observed-plus-reported with reported-only scoring are the principal mitigations. The RFPI is newly constructed and its measurement properties are not yet established; internal consistency, inter-rater reliability and structural properties will be reported and the primary result interpreted in light of them. With 18 clusters, the trial has limited degrees of freedom at the cluster level, which constrains covariate adjustment and gives limited precision for the ICC estimate; the Kenward–Roger correction and a cluster-level robustness analysis are prespecified in response, and adequate power depends on the baseline-adjusted analysis. Cluster heterogeneity across a large county may be substantial, and stratification by sub-county reduces but does not eliminate this. Contamination cannot be excluded, since CHPs and caregivers move between communities; it will be measured and examined in a sensitivity analysis. The three-month endline captures proximal behaviour change and is not designed to detect effects on child growth. Finally, the exclusion of caregivers under 18 years limits generalisability to adolescent mothers.

## Conclusion

This trial will generate evidence on whether a structured, competency-based responsive feeding package delivered by Community Health Promoters improves caregiver responsive feeding practices in an arid and semi-arid Kenyan setting, and on whether that package can be delivered with acceptable reach, adoption and fidelity within routine community health services. By combining a randomized effectiveness comparison with a prospectively specified implementation evaluation, the trial is intended to inform decisions about integrating responsive feeding counselling into the Kenyan community health platform and about its relevance to comparable pastoral and ASAL contexts.

## Declarations

### Trial registration

Registry: Pan African Clinical Trials Registry (PACTR). Registration number: PACTR202611787801603. Date of registration: 9 November 2026. The trial was registered prospectively; first enrolment is planned for 11 January 2027. The registry record was updated on 15 November 2026 to reflect protocol version 1.1.

### Ethics approval and consent to participate

The trial has been approved by the Masinde Muliro University of Science and Technology Institutional Scientific and Ethics Review Committee (MMUST/ISERC/314/2026, approved 17 August 2026) and licensed by the National Commission for Science, Technology and Innovation (NACOSTI/P/26/4195285, 14 September 2026). Authorisation from the West Pokot County Department of Health will be obtained before enrolment. Written informed consent will be obtained from all participants.

### Author contributions

Conceptualization: AKK, ENM, SOK. Methodology: AKK, ENM, SOK, MSS. Investigation: AKK. Project administration: AKK. Data curation: AKK. Validation: ENM, SOK, MSS. Supervision: ENM, SOK. Writing – original draft: AKK. Writing – review & editing: AKK, ENM, SOK, MSS.

### Roles and responsibilities

AKK is the principal investigator and is responsible for trial conduct, data integrity and day-to-day management. ENM and SOK are the academic supervisors of the doctoral work and provide scientific oversight. MSS is the corresponding author. The allocation sequence will be generated by an independent statistician who is not an author and has no other role in the trial. The primary sponsor is the principal investigator (AKK), as an individual investigator-sponsor; sponsor contact details are given in the PACTR record. There is no secondary sponsor, and no sponsor or funder holds authority over study design, data collection, analysis, interpretation, or the decision to publish.

## Acknowledgements

The authors acknowledge the West Pokot County Department of Health, the Kacheliba and Kipkomo sub-county health management teams, Community Health Unit committees, Community Health Promoters and the caregivers who will take part in this study. A generative artificial intelligence tool (Claude, Anthropic) was used for language editing and consistency checks; the authors reviewed all content and take full responsibility for it.

## Supporting information

**S1 File. SPIRIT 2025 checklist.**

**S2 File. SPIRIT-Children and Adolescents (SPIRIT-C) 2026 checklist.**

**S3 File. Ethics-approved study protocol.** Doctoral research proposal approved by MMUST-ISERC (MMUST/ISERC/314/2026), including the study instruments, participant information sheet and consent form, with the ethics approval letter and NACOSTI research licence.

**S4 File. WHO Trial Registration Data Set (version 1.3.1) record for PACTR202611787801603**

